# Face masks to prevent transmission of respiratory diseases: Systematic review and meta-analysis of randomized controlled trials^*^

**DOI:** 10.1101/2020.07.31.20166116

**Authors:** Hanna M. Ollila, Markku Partinen, Jukka Koskela, Riikka Savolainen, Anna Rotkirch, Liisa T. Laine

## Abstract

**Objective:** To examine the effect of face mask intervention in respiratory infections across different exposure settings and age groups.

**Design:** Systematic review and meta-analysis.

**Data sources:** PubMed, Cochrane Central Register of Controlled Trials, and Web of Science were searched for randomized controlled trials investigating the effect of face masks on respiratory infections published by November 18th, 2020. We followed PRISMA guidelines.

**Eligibility criteria for selecting studies:** Randomized controlled trials investigating face masks in respiratory infections across different exposure settings. Two reviewers performed the search, extracted data, and assessed the risk of bias. Random effects meta-analysis with risk ratio, adjusted odds ratios, and number needed to treat were performed. Findings by source control or wearer protection, age groups, exposure settings, and role of non-compliance were evaluated.

**Results:** Seventeen studies were included, (N=11,601 cases and N=10,286 controls, follow-up from 4 days to 19 months). Fourteen trials included adults and children and three trials included children only. Twelve studies showed non-compliance in treatment and eleven in control group. Four studies supported the use of face masks. Meta-analysis across all studies with risk ratios found no association with number of infections (RR=0.957 [0.810 − 1.131], p=0.608). Meta-analysis using odds ratios adjusted for age, sex, and vaccination (when available) showed protective effect of face masks (OR=0.850 [0.736 − 0.982], p=0.027). Subgroup meta-analysis with adjusted odds ratios found a decrease in respiratory infections among adults (14 studies, OR = 0.829 [0.709 − 0.969], p=0.019) in source control setting (OR = 0.845 [0.7375 − 0.969], p=0.0159) and when face masks were used together with hand hygiene OR = 0.690 [0.568 − 0.838], p=0.0002). Overall between-study heterogeneity was large also in the subgroup analyses.

**Conclusion:** Despite the large between study heterogeneity, compliance bias and differences by environmental settings, the findings support the use of face masks to prevent respiratory infections. *PROSPERO registration number CRD42020205523*.

## 1. Introduction

The COVID-19 and other pandemics are a scourge causing severe losses on health, economy, and well-being [1, 2]. COVID-19, caused by SARS-CoV-2 (Severe Acute Respiratory Syndrome Coronavirus 2), can spread through droplet-mediated transmission through contaminated surfaces and air [3–6]. Non-pharmaceutical interventions (NPIs), such as maintaining physical distance, appropriate hand hygiene, and face masks have been adopted as the primary tools to limit the number of COVID-19 infections [7] while the vaccines are being developed and pharmaceuticals are studied for repurposing.

Prior to COVID-19 pandemic, the use of face masks by the general public was considered a relatively new policy tool in preventing person-to-person transmission on a global scale. Face masks are widely used in health care settings. Prediction models suggest that universal use of face masks in public may have a substantial preventive impact on disease spread, even without medical masks or 100% compliance [8–10]. In addition, a pooled meta-analysis of the spread of infectious viral diseases of up to 172 studies showed a consistent effect regarding the efficacy of face masks in preventing infections by SARS-CoV-2 and the betacoronaviruses that cause severe acute respiratory syndrome, and Middle East respiratory syndrome [7].

However, the evidence on the efficacy of face masks use among the general public from randomized controlled trials has been noted as being only suggestive [11]. For example, many of the randomized controlled trials have documented non-compliance either in the face mask intervention arm [12–23] or in the control arm [12, 15, 17, 18, 21, 23–28]. Because these studies estimate the intention-to-treat effect of face masks, non-compliance can underestimate the magnitude of the treatment effect of face masks use for a given randomized controlled trial.

The aim of this systematic review and meta-analysis was to examine the evidence from randomized controlled trials of face masks in the context of COVID-19 or diseases which spread through similar mechanisms to COVID-19: respiratory infections. An earlier systematic review and meta-analysis has investigated the effect of face masks by focusing on the use of cloth masks [29] in non-health care settings while [30] combined various types of studies, including randomized controlled trials, case-control studies and cohort studies. Our review complements these studies by focusing solely on randomized controlled trials in different exposure settings (hospital, household, and community) and age groups (adults vs. children). Moreover, we examine the role of non-compliance in treatment and control arms, study whether the findings are affected by source control or wearer protection, and the role of hand hygiene guidance on face masks.

## 2. Methods

This systematic review was performed according to the Preferred Reporting Items for Systematic Reviews and Meta-Analyses [31]. Our review protocol was registered on PROSPERO in November 2020 (registration number CRD42020205523).

### 2.1 Search strategy

We performed the searches using the Cochrane Central Register of Controlled Trials (CENTRAL), PubMed, and Web of Science (science and social science citation index). We performed the PubMed search using Medical Subject Headings (MeSH) listed in Supplement A. In other search engines, we used the following search terms: facemasks/face masks AND/OR infection. The full search protocol with the criteria are described in Supplement A. The searches were limited to randomized controlled trials on humans published by November 18th, 2020. We did not limit the searches by language. The search results were uploaded on Endnote, and the unique citations were kept and screened.

### 2.2 Study selection, inclusion and exclusion criteria

We included randomized controlled trials on humans (general population and health care personnel in a risk of contracting respiratory infectious diseases) that compared face mask use (FFP1, FFP2, FFP3, cloth mask or surgical mask) or face mask and hand hygiene or face mask and education with no face mask use. We did not make exclusions based on a setting, instead, we included interventions that were executed in various settings, such as in health care, community, or household.

We included two summary statistics measuring the risk of infection to our analysis. First is the unadjusted relative risk for infection which we computed using the raw numbers of infected in treatment arms which were provided in the studies. However, it is possible that omitted variables such as age, sex, vaccination or other factors such as correlation of outcomes within household affect the outcomes. Because almost none of the studies provided relative risk estimates adjusted for covariates, we used adjusted odds ratios in our secondary analyses if they were available in the studies. We discuss the limitations of these different summary statistics (odds ratio, risk ratio, for example) used in meta-analyses in detail in Section 4.3.

We excluded interventions that compared different types of face masks to each other (in which the comparison arm were assigned to use a face mask). We did not exclude any studies based on age and gender or have exclusion criteria based on sample sizes or follow-up periods. We included all the studies with a whole text available (including pre-prints) while we excluded the studies which had only an abstract available. Table A1 in Supplement A provides a detailed summary of the inclusion and exclusion criteria.

Two authors (HMO and LTL) executed the search. The authors (HMO and LTL) independently reviewed all titles and abstracts to define the papers that could potentially be included in the systematic review. After this, both authors independently screened the articles and determined whether they met the inclusion and exclusion criteria. The disagreements between the two authors were resolved by discussion.

### 2.3 Data extraction

Two authors (HMO and LTL) independently extracted the data which included (1) study setting (time, country, population); (2) intervention details (randomization level, follow-up, type of mask, other interventions, case or index case definition); (3) outcome measures (N per arm and odds ratio effect sizes with respective standard errors); (4) compliance measure; and (5) study results for the effects of face mask use. Two other reviewers (JK and RS) checked the extracted data for errors. We contacted the corresponding author of [17] to confirm our interpretation of their data and also asked if they were able to provide the raw numbers for source control estimates. Unfortunately the data was not available anymore.

### 2.4 Risk-of-bias assessment

Two review authors (HMO and LTL) independently assessed the risk of bias using the Cochrane Risk of Bias tool [32]. Any discrepancies or unusual patterns were resolved by consensus. The following characteristics were evaluated: Random sequence generation, allocation concealment (selection bias), blinding of participants and personnel (performance bias), blinding of outcome assessment (detection bias), incomplete outcome data (attrition bias), selective reporting (reporting bias), and non-compliance in the treatment arm and control arm. The risks were categorized as low risk, unclear risk or high risk of bias. Following the Cochrane tool for risk assessment, we denoted the overall risk of bias as low if all the categories were at a low risk of bias, high if at least one domain was at a high risk of bias. We denoted the overall risk of bias as unclear if at least one domain was at an unclear risk of bias and no domain was at a high risk. Taking the overall risk into account is important because it helps in avoiding the bias caused by prioritizing one category over others as any source of bias can be problematic [32].

### 2.5 Data analysis

The results for all the outcomes were expressed as risk ratios (RR), odds ratios (OR), and 95% confidence intervals for the effect estimates. We combined the estimates using a random-effects meta-analysis, based on the assumption that the existence of methodological and clinical heterogeneity potentially affecting the results was likely. We estimated the between-study variance by using the DerSimonian and Laird method of moments -estimator. We calculated the 95 percent confidence intervals using the Wald method.

We assessed heterogeneity and quantified statistical inconsistency by using the 2 test and the I2 statistic, respectively [33]. We used stratified meta-analyses to explore heterogeneity in the effect estimates according to: source control and wearer protection, non-compliance, study populations, and settings. We studied how non-compliance in controls (using a face mask) is associated with the intervention effects in the meta-analysis with a meta-regression.

The small study effects were studied by generating contour-enhanced funnel plots to examine the bias in the results of the meta-analysis (the tendency for intervention effects from smaller studies to differ from those estimated in larger ones, which can result from reporting biases, methodological or clinical heterogeneity or other factors).

We conducted all the analyses using the meta, metaphor, and dmetar packages in R version 4.02 and meta package in Stata version 16.

### Patient and public involvement

Our study research was not informed by patient and public involvement because we use secondary data. Patients or the public were not involved in the design, conduct, reporting or dissemination plans of our research.

## 3 Results

### 3.1 Search results

Our search resulted in 2,354 unique publications. After the review, we retained 17 randomized controlled trials of face mask use while 2,337 articles were excluded because they did not meet the inclusion criteria (Figure 1).

**Figure 1.**
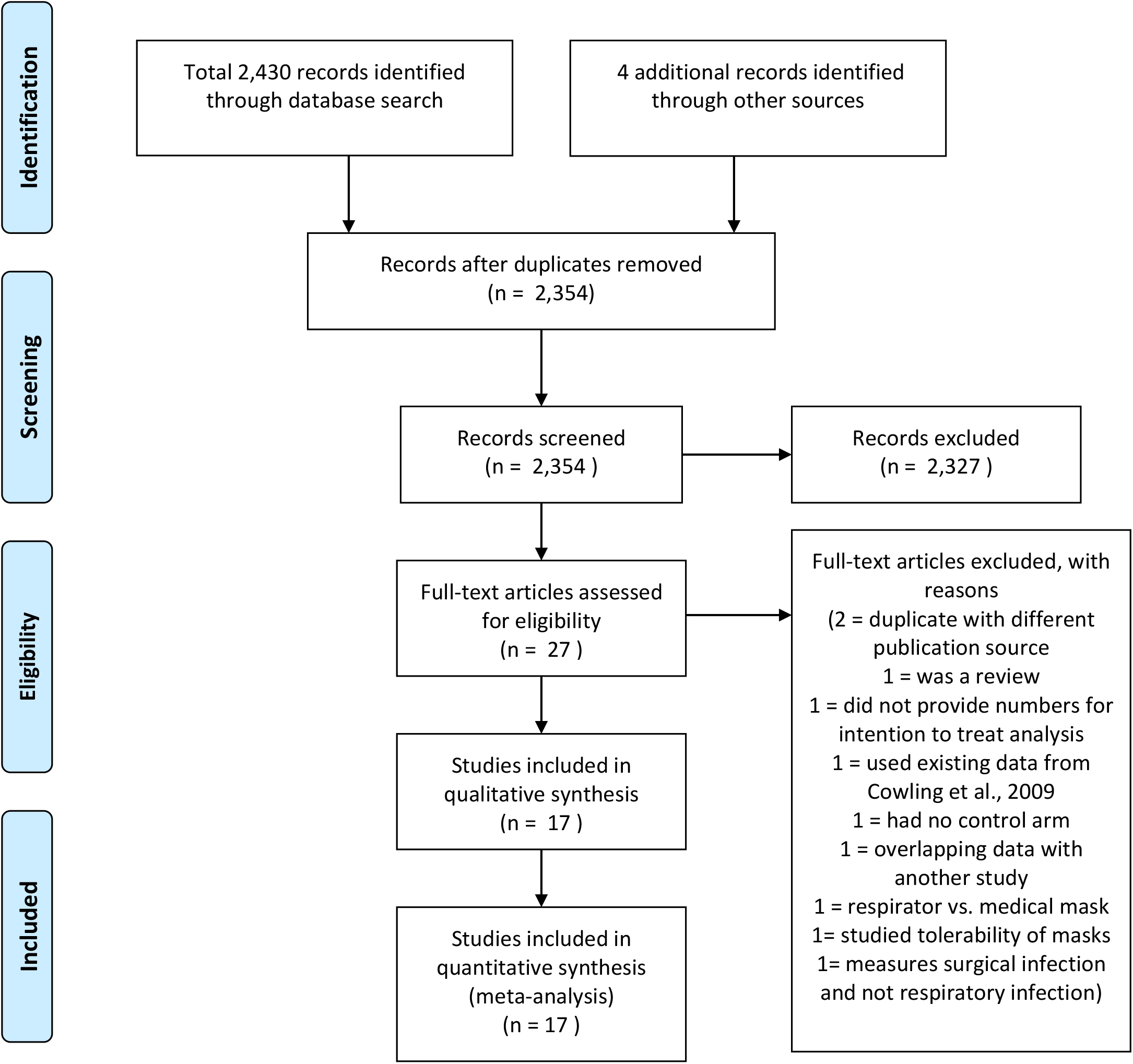
Prisma flow diagram of included articles.

The studies included a variety of environmental settings: pilgrims (3 studies), college students (2 studies), healthcare (4 studies) to household environment (7 studies). Six trials were performed in a community setting [12–16, 24]. Three included children only [20, 22, 23] and 14 trials included to both adults and children [12–19, 21, 24–28].

### 3.2 Characteristics of included studies

Supplementary table 2 summarizes the characteristics of each study. Altogether, these studies included 11,601 study participants in the treatment arm and 10,286 in the control arm. The duration of follow-up varied from 4 days to 19 months. Nine of the seventeen studies reported raw values for wearer protection estimates exclusively [13–16]. Seven of the seventeen studies reported raw values for source control estimates exclusively [17–20, 22–25]. For study [19][19] raw values for source control were not available.

Studies were carried out in eleven countries in several continents: Australia [20], China [21, 27], Denmark [16], France [25], Germany [22], Hong Kong [17, 18], Japan [26], Saudi Arabia [12, 15, 24],Thailand [23], the United States [13, 14, 19], and Vietnam [28].

### 3.3 Assessment of intervention: face mask use

In addition to conducting interventions in diverse settings (community, hospital, household) and age groups (adults, children), the interventions themselves varied. In some of the interventions, the treatment arm received an education leaflet in addition to face masks [12], while, in others, the intervention included a weekly supply of face masks and a plastic bag for storage and daily disposal [15]. The type of face mask varied from cloth masks to medical masks with ear loops. Some trials had a separate hand hygiene, face mask or hand hygiene with face mask arm [13, 14, 18, 19] in which the treatment arm the intervention included also a hand sanitizer or instruction to wash hands.

Three studies [13, 14, 24] found a protective effect of face masks in the intention-to-treat analysis and one in the analysis of secondary transmission rate [19]. Two of these studies had a follow-up length of 6 weeks [13, 14] and third one up to 19 months [19]. These three studies [13, 14, 19] had also the longest follow-up times among the 17 included interventions. In addition, two additional studies showed an association in the per protocol analysis [18, 34]. These studies were source control studies where an early intervention within 36h was associated with a reduced number of respiratory infections in the contacts, suggesting that face masks may be most efficient if adopted early on during the exposure so that transmission and infection has not already happened before adopting mask use.

### 3.4 Risk of bias across the studies

Table 1 summarizes the risk of bias on the study level. The observed bias was low or unclear in the majority of the 17. As discussed in our methods section, we did not exclude studies based on bias. In the instances in which a bias was found, the main concerns were related to non-compliance either in the treatment (12 studies [12–23]) or in the control arm so that treated individuals did not use the mask while individuals in the control arm did use it (11 studies [12, 15, 17, 18, 21, 23–28]). Almost all the trials had an increased a risk of bias due to unclear or a lack of blinding. Obviously, blinding per mask use is challenging due to the visible nature of face mask. In addition, one study could not allocate the control arm randomly due to local health regulations, so it recruited a separate (non-randomized) control arm and examined primarily differences between face masks [27]. There were some concerns due to the lack of blinding at the stage of identification of symptoms per treatment arm (13 studies [12–19, 21, 24, 26–28]). Similarly, it is unclear if the outcome assessment was fully blinded in any of the 17 studies (Table 1).

**Table 1.** Bias assessment. Non-compliance in the treatment or control group: high risk if the reported non-compliance was greater than 50%, unclear if between 30-50%, and low if under 30%.

|  | Sequence generation | Allocation concealment | Blinding |  | Incomplete outcome data | Selective reporting | Non-compliance |  |
| --- | --- | --- | --- | --- | --- | --- | --- | --- |
|  |  |  | Participants and personnell | Outcome assesment |  |  | Treatment group | Control group |
| Abdin et al., 2005 | Unclear risk | Unclear risk | Unclear risk | Unclear risk | Low risk | Low risk | Unclear risk | Unclear risk |
| Aiello et al., 2010 | Unclear risk | Low risk | Unclear risk | Unclear risk | Low risk | Low risk | Unclear risk | Low risk |
| Aiello et al., 2012 | Low risk | Low risk | Unclear risk | Unclear risk | Low risk | Low risk | Unclear risk | Low risk |
| Alfelali et al., 2020 | Unclear risk | Unclear risk | Unclear risk | Unclear risk | Low risk | Low risk | Unclear risk | High risk |
| Barasheed et al., 2014 | Low risk | Low risk | Unclear risk | Unclear risk | Low risk | Low risk | Low risk | Unclear risk |
| Bungaard et al. 2020 | Low risk | Low risk | Unclear risk | Unclear risk | Low risk | Low risk | Unclear risk | Low risk |
| Canini et al., 2010 | Low risk | Low risk | Low risk | Unclear risk | Low risk | Low risk | Low risk | Unclear risk |
| Cowling et al., 2008 | Low risk | Low risk | Unclear risk | Unclear risk | Low risk | Low risk | Unclear risk | Unclear risk |
| Cowling et al., 2009 | Low risk | Low risk | Unclear risk | Unclear risk | Low risk | Low risk | Unclear risk | Unclear risk |
| Jacobs et al., 2009 | Low risk | Low risk | Unclear risk | Unclear risk | Low risk | Low risk | Low risk | Unclear risk |
| Larson et al., 2010 | Low risk | Low risk | Unclear risk | Unclear risk | Low risk | Low risk | Unclear risk | Low risk |
| Macintyre et al., 2009 | Low risk | Low risk | Low risk | Unclear risk | Low risk | Low risk | Unclear risk | Low risk |
| Macintyre et al., 2011 | High risk | Unclear risk | Unclear risk | Unclear risk | Low risk | Low risk | Low risk | Unclear risk |
| Macintyre et al., 2015 | Low risk | Unclear risk | Unclear risk | Unclear risk | Low risk | Low risk | Low risk | Unclear risk |
| MacIntyre et al., 2016 | Low risk | Low risk | Unclear risk | Unclear risk | Low risk | Low risk | Unclear risk | Unclear risk |
| Simmerman et al., 2011 | Low risk | Low risk | Low risk | Unclear risk | Low risk | Low risk | High risk | Unclear risk |
| Suess et al., 2012 | Low risk | Low risk | Low risk | Unclear risk | Low risk | Low risk | High risk | Low risk |

**Table 2.**
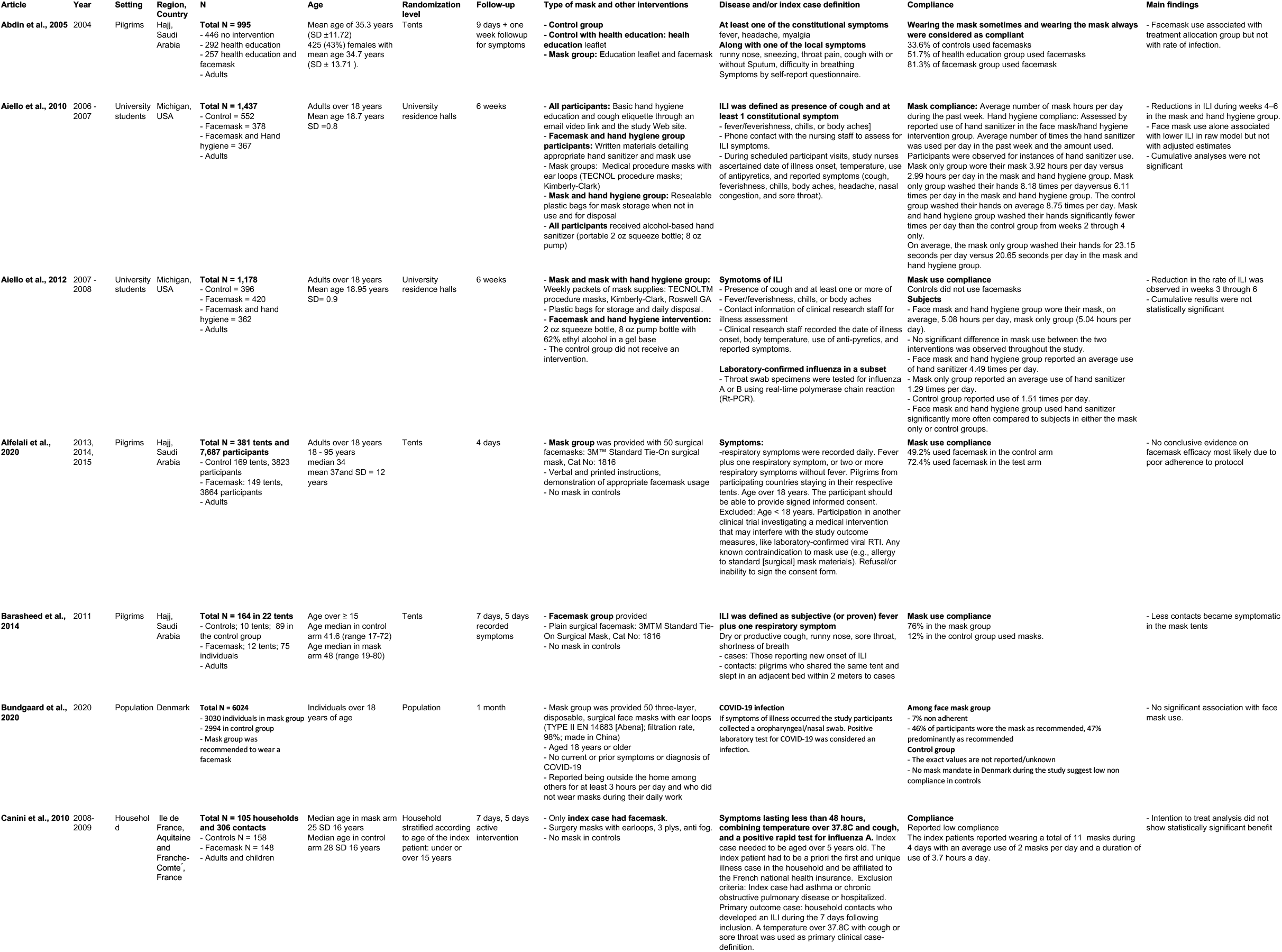
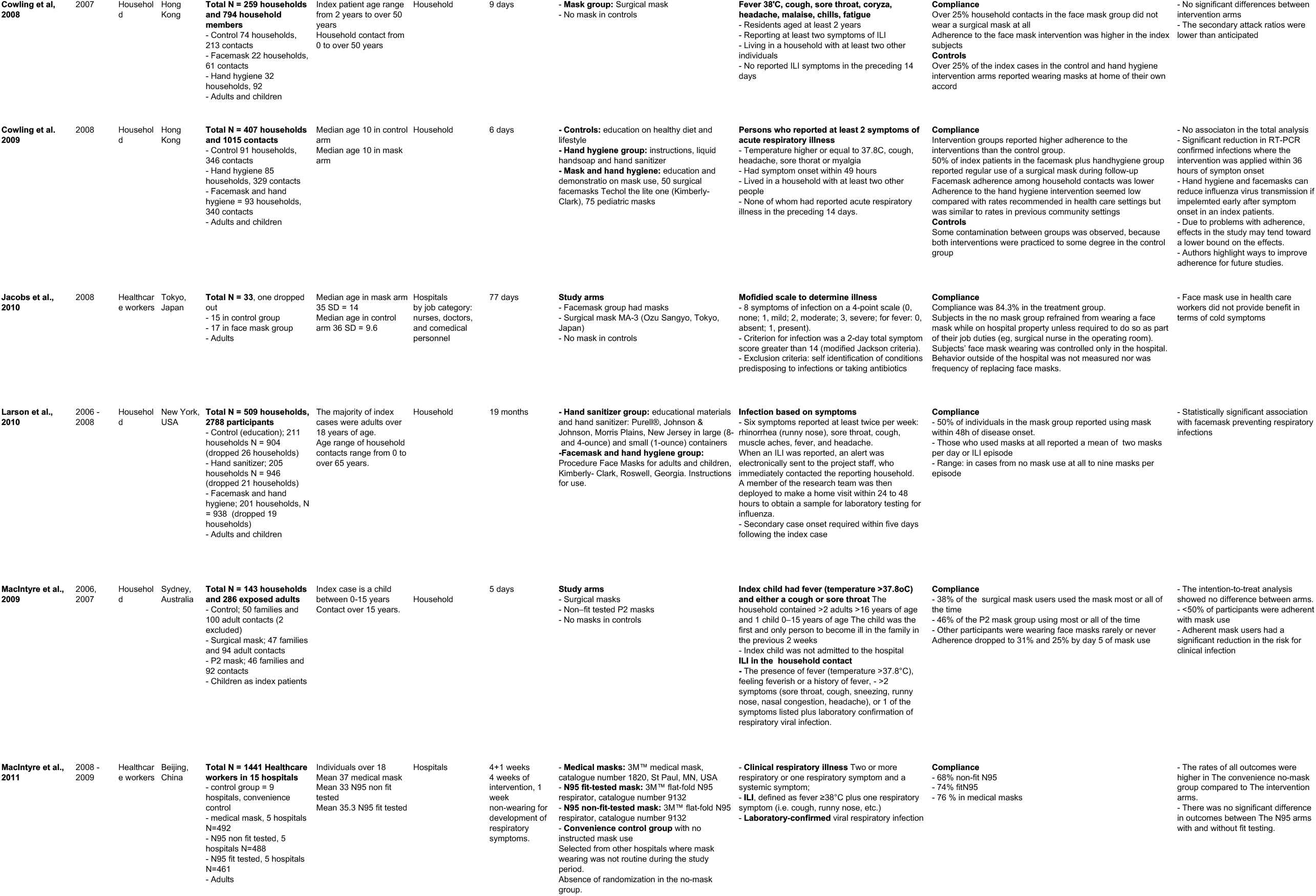
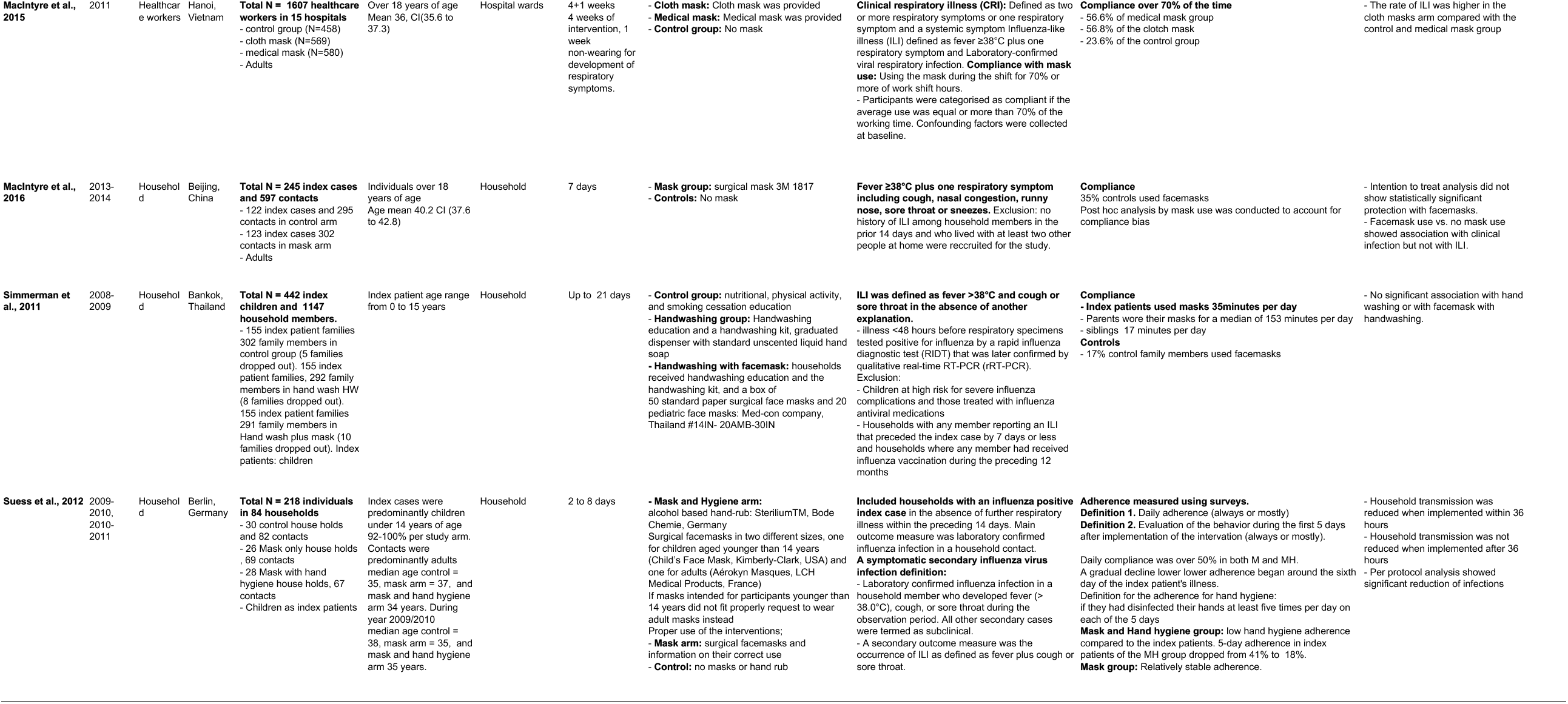
Characteristics of included studies.

### 3.5 Details about random sequence generation and allocation concealment were unclear for some trials

A summary of the proportion of the trials that were at low, unclear, and high bias for each domain is shown in (Figure 2). We found no evidence of a publication bias by a visual examination of funnel plots (Figure 7) or by an analysis based on Egger’s tests: *β* = 0.36, se = 0.52, p = 0.496.

**Figure 2.**
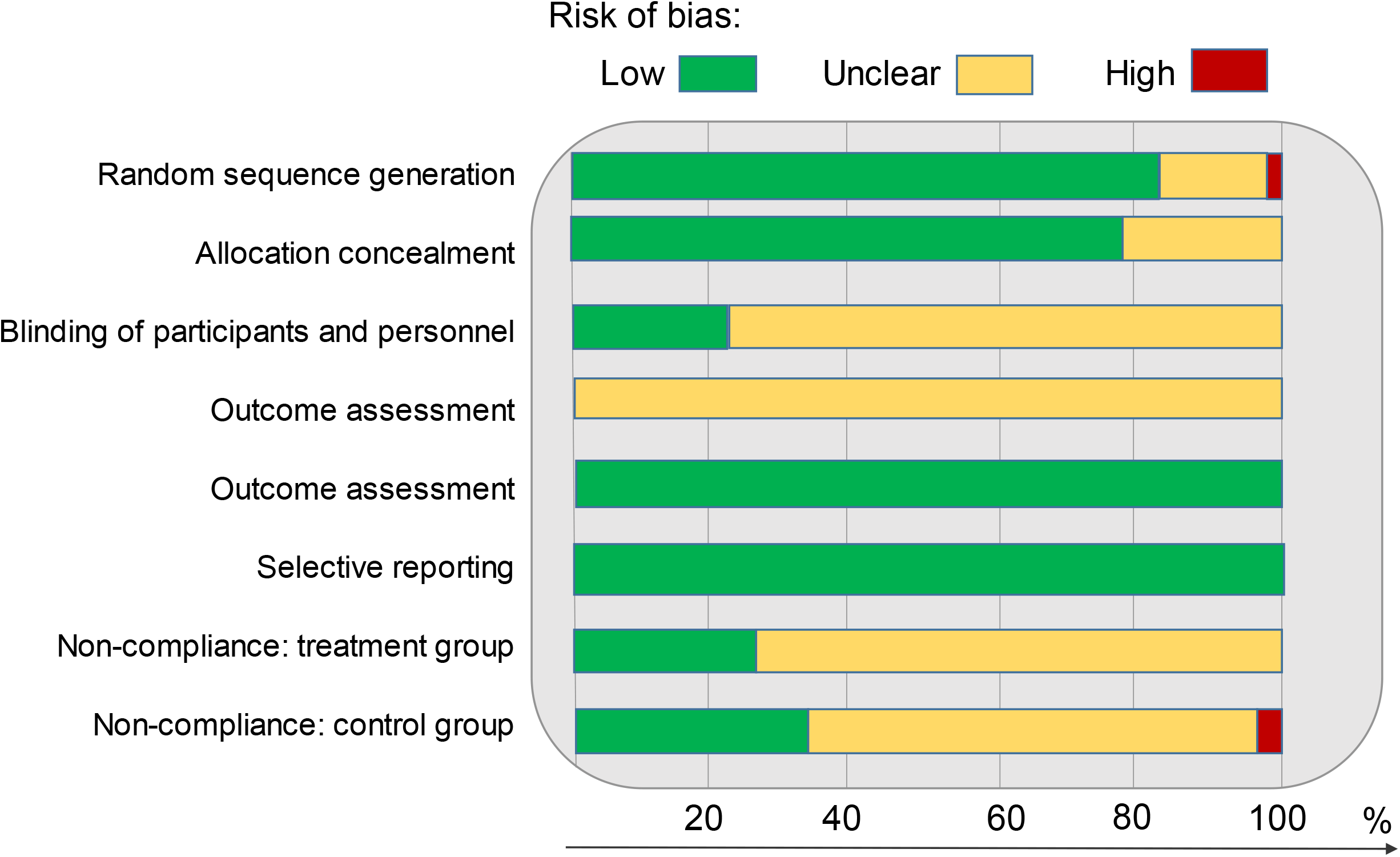
Review authors’ judgement of each of risk-of-bias item as percentage across the included studies.

### 3.6 Face masks and respiratory infections

There were in total 1,542 events among the treatment arms (N total treatment = 11, 601) and 1, 345 events in control arms (N total control = 10, 286). The meta-analysis using unadjusted risk ratios (from raw N values) across all 17 studies finds no association between face mask intervention and respiratory infections (RR = 0.957 [0.810 − 1.131], p = 0.6086) (Figure 3a). The analysis using covariate adjusted odds ratios shows a protective effect of face masks (OR = 0.850 [0.736 − 0.982] p = 0.027, Figure 3b).

**Figure 3.**
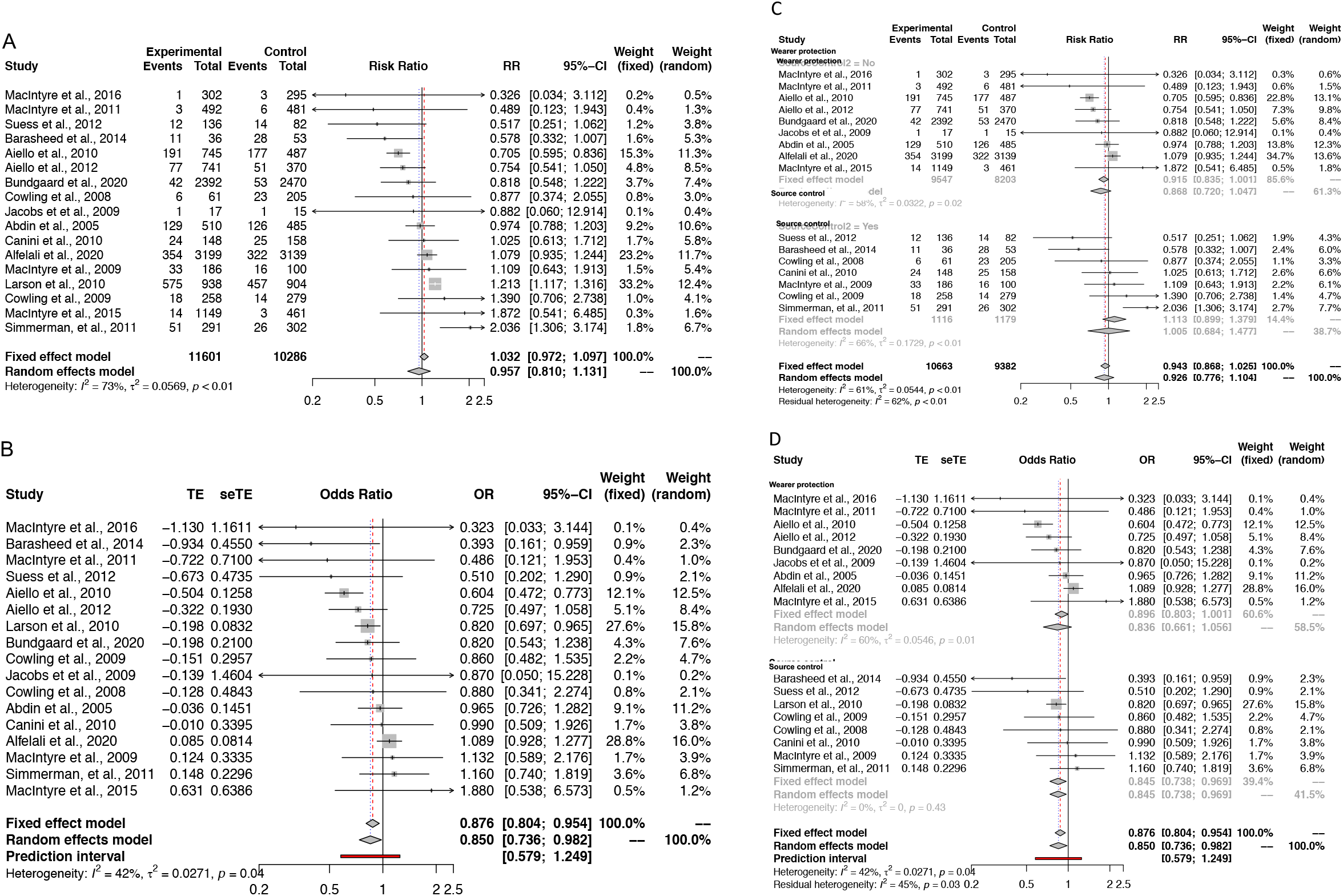
Pooled effect across all 17 studies using a) raw estimates and relative risk b) adjusted odds ratios when available c) raw estimates examining source control and wearer protection separately and d) adjusted odds ratios examining source control and wearer protection separately. TE = treatment effect and seTE = standard error of treatment effect. Fixed and Random effects pooled estimates are shown.

The overall effect of face masks consists of the combination of source control and personal protection for the mask wearer (wearer protection). We investigated this mechanism by performing a subgroup analysis by categorizing the studies exclusively based on their estimated effects on source control or wearer protection. Subgroup meta-analysis based on the unadjusted risk ratios was inconclusive (wearer protection RR = 0.868 [0.720 − 1.047], p = 0.139; source control RR = 1.0054 [0.685−1.477], p = 0.978, Figure 3c). Secondary analysis using adjusted odds ratios was also inconclusive on wearer protection (OR = 0.836 [0.661 − 1.056] p = 0.133) but indicates a protective effect as a source control (OR = 0.845 [0.738 − 0.969] Figure 3d). Similarly, raw values did not associate with fewer infections in individuals over 15 years of age (RR = 0.920 [0.774 − 1.094], p = 0.343, Figure 4) whereas adjusted ORs did (RR = 0.829 [0.709 − 0.969] p = 0.019).

**Figure 4.**
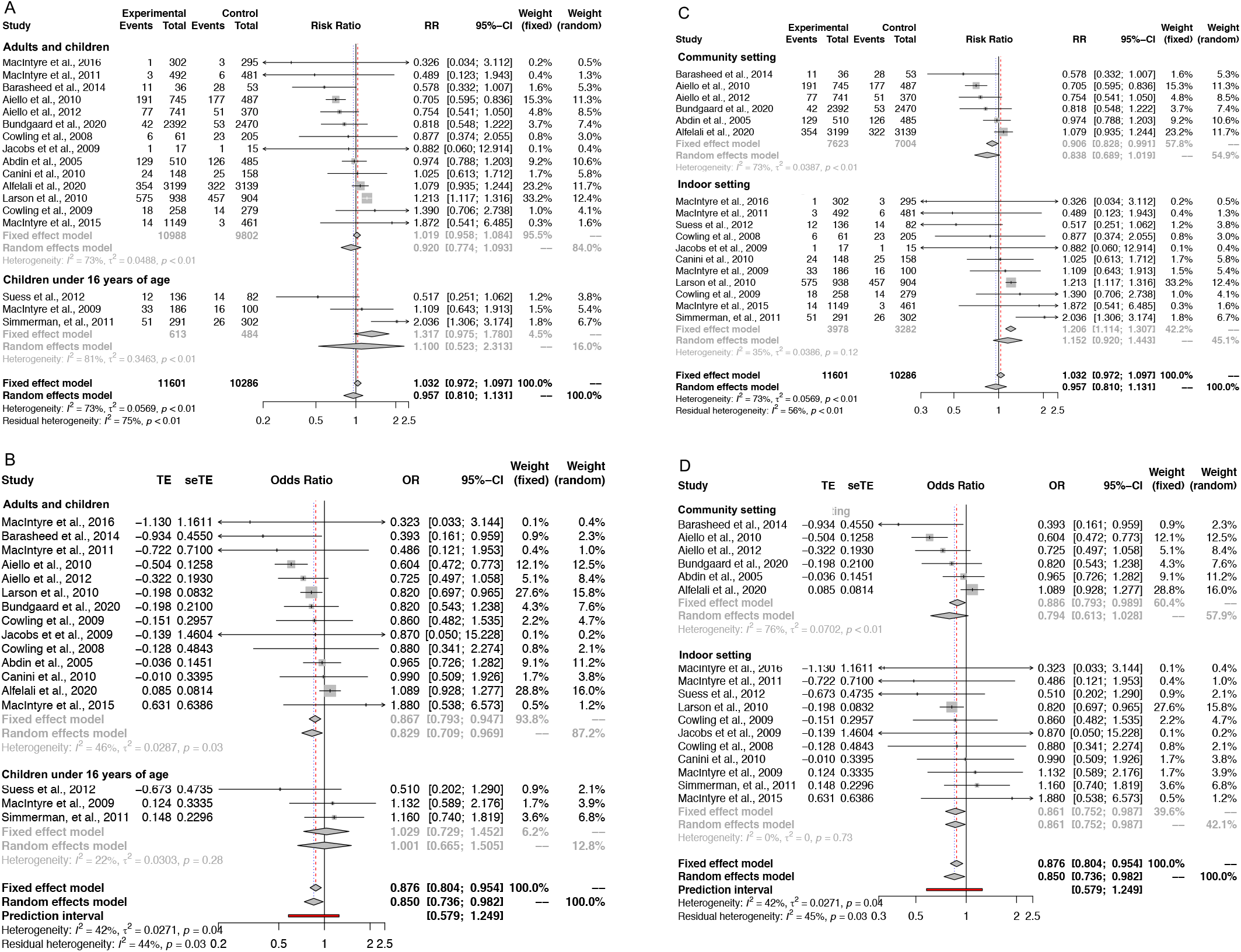
Subgroup analysis in a) adults and in children using raw estimates and relative risk b) adults and in children using adjusted odds ratios when available c) community setting and indoor environment separately using raw estimates and d) adjusted odds ratios examining community setting vs indoor environment using raw estimates separately. TE = treatment effect and seTE = standard error of treatment effect. Fixed and Random effects pooled estimates are shown.

In addition to the substantial differences between the analysis using unadjusted and adjusted effect estimates, it is also possible that individual studies bias the estimates of the meta-analysis. Thus, we performed a meta-analysis through a leave-one-out analysis to examine if one study causes a systematic association to a given direction. The effect sizes were all systematically at RR < 1. However, one of the largest studies, [15], had a significant level of non-compliance in the treatment arm with 49% of the controls using face masks; and the second largest study which had the longest follow-up time, [19], we did not have access to secondary transmission values.

We investigated the importance of non-compliance on the results by performing a subgroup meta-analysis without the studies with non-compliance of over 10% in the control arm. This showed a protective effect of face masks in adjusted ORs but not with raw estimates (RR = 0.877 [0.686 − 1.121], OR = 0.7641 [0.682 − 0.856] p < 0.0001) indicating that non-compliance has weakened the power to observe an association in these trials.

Environments differ by their risk of contracting respiratory viruses due to having varying amount of viral particles or a different length of exposure. As a result, effects of face masks likely differ by the length and the setting of the exposure. We investigated potential differences by conducting a subgroup meta-analysis of different environments: community and hospital or household settings by focusing on studies that included adults (Figure 4). In the random effects meta-analysis in the hospital or household settings association was seen with adjusted ORs but not with raw effect estimates (RR = 1.207 [1.114 − 1.307], p = 0.219, OR = 0.861 [0.752 − 0.987], p = 0.0318). The effect was similar in the community settings although statistically insignificant (RR = 0.838 [0.689 − 1.012], p = 0.077, Figure 4). It is possible that these large confidence intervals in the community setting result from non-compliance (between 10% and 50% of the control groups using masks) in three out of the six studies that assessed community transmission and from relatively low compliance in the treatment groups. Indeed, meta-regression adjusting for face mask use in the control arm showed statistically significant association at the population setting (RR = 0.86 [0.79-0.94], p = 0.0007).

In the twelve of the trials including adults, the intervention consisted solely of face mask use while in six the intervention included also guidance on appropriate hand hygiene together with the face mask use. This subgroup analysis for the face mask with hand hygiene guidance showed effect with adjusted ORs but not with raw estimates (RR = 1.020 [0.738 − 1.408], p = 0.900, OR =0.690 [0.568 − 0.838], p = 0.0002, Figure 5).

**Figure 5.**
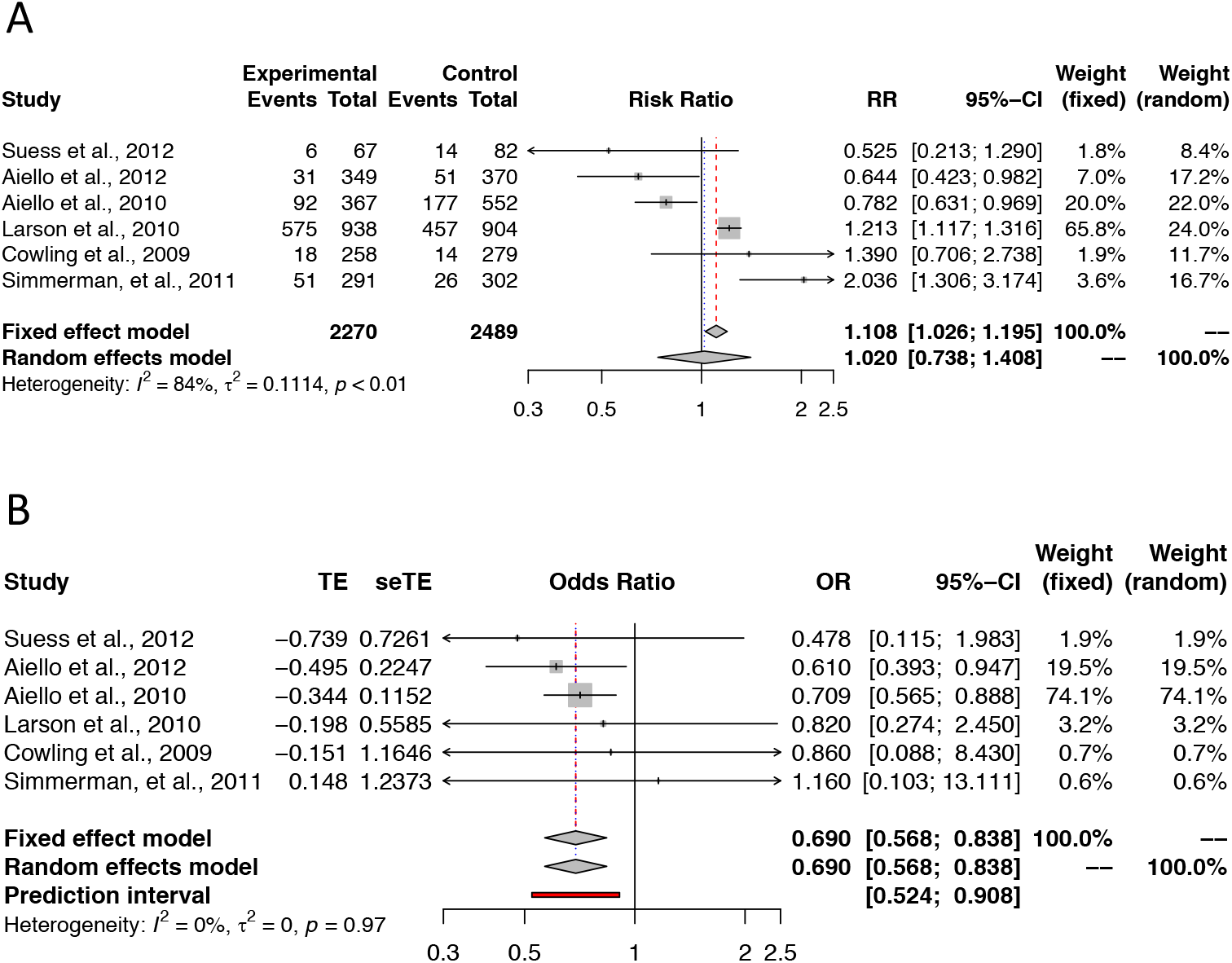
Subgroup analysis in a) face mask together with instructed hand hygiene using raw estimates b)) face mask together with instructed hand hygiene using adjusted odds ratios when available. TE = treatment effect and seTE = standard error of treatment effect. Fixed and Random effects pooled estimates are shown.

### 3.7 Number needed to treat

We approximate the effect of masks on population health by exploring the number needed to treat (NNT), that is, how many individuals need to wear a mask to prevent one person from contracting a respiratory infection. The number needed to treat depends on such infections at the population level. If there are few infections, a larger number of mask users will be needed to prevent one infection. Based on the results from this meta-analysis and assuming a low baseline risk of 0.01, NNT is 455. With a larger baseline risk, 0.05, NNT is 91, and with a higher still risk, 0.2, NNT becomes 23.

## 4 Discussion

### 4.1 Main findings

This systematic review and meta-analysis of 17 randomized controlled trials examined whether face mask intervention can prevent respiratory infections. 4 out of 17 studies supported the use of masks. In addition, the analysis of the adjusted odd ratios showed a protective effect of face masks across different settings. However, the pooled effect using the unadjusted effects based on raw numbers versus adjusted estimates were inconsistent. This together with remaining heterogeneity even in subgroup analyses indicate large variation across these studies that needs to be taken into account both in the interpretations of the results and when designing future research on face masks. The protective effect of face masks was observed both with face mask use alone and when face masks were combined with appropriate hand hygiene. This result is aligned with the current evidence that face masks are most efficient when used together with appropriate hand hygiene and other protective measures. It is worth noting that, despite the relatively large between-study heterogeneity and small effect sizes in the individual studies, the findings did support use of face masks. Therefore, these findings together with the mounting other evidence suggest that face masks may be considered as a useful NPI for respiratory infections, including COVID-19.

### 4.2 Quality of evidence

We found 17 randomized controlled trials that had assessed whether masks affect the number of respiratory infections. Other earlier studies have been conducted using case-control settings or with masks with a strong filtering capacity [7]. Earlier systematic reviews and meta-analyses have investigated, for example, the effect of face masks by combining types of studies, including randomized controlled trials, case-control studies and cohort studies [30] or cloth masks [29] in non-healthcare settings. The findings from our systematic review and meta-analysis are in line with the conclusions from these earlier meta-analyses conducted in different settings. In contrast, by including as a full set of studies as possible, we are better powered in estimating the effect of a mask intervention.

While the intention-to-treat analysis yields an unbiased estimate of the efficacy of the face mask intervention, its magnitude is biased downwards relative to the actual treatment effect of face masks. While the overall quality of the earlier trials is solid, there were biasing factors across the studies, including a compliance bias either because of low compliance in the face mask treatment arm or the use of face masks in the control arm, which may bias the estimates towards the null hypothesis. This is also confirmed by our sensitivity analysis.

In addition, as the effects with hand hygiene seems to be stronger than with mask use alone, one might conclude that hand hygiene is driving the association while mere face masks do not protect from respiratory infections. Indeed, while masks have been shown to be effective in themselves, their impact and, therefore, efficacy is largest when combined with other protective measures [7]. Also in our study, the effect of masks was further accentuated when combined with complementary measures, such as improved hand hygiene [13, 14]. Furthermore, other complementary measures for disease control, such as physical distancing measures, have an impact on the spread of diseases and the number of particles in the air and, hence, also add to the effect of face masks.

Indeed, in a review [35], the estimated number needed to mask to prevent one infection ranged from three (N95 masks) to six (face masks), and the number is higher still when the infection risk is low to start with. Clearly, these NNTs are only approximations since the reproduction number R differs between viral infections. Similarly, if there are no active infections, the NNT will be infinite: no infections can be prevented as none are present in the population.

With these limitations in mind, we calculated that, for respiratory infections, the NNT might range from 23 to 455. To put this into context, let us presume that, in a metropolitan area with a population of one million, 30% of the residents use face masks. With NNT=455, this might prevent 600 respiratory infections. This effect size is comparable to the NNT of pharmaceuticals. For example, the NNT for statin, one of the most widely prescribed drugs, in primary prevention of major vascular events at low levels of a CVD risk (5-10% within 5 years) ranges from 67 to 170 and is of a similar scale to face masks [36].

We show that the studies where hand hygiene was assessed together with mask use, effects with multiplicative protective measures were seen. Our results support use of face masks in preventing respiratory infections and, hence, the WHO guidelines that recommend the use of face masks together with physical distancing and hand hygiene in controlling the spread of COVID-19.

### 4.3 Limitations

First of all, the populations studied here had residual heterogeneity. Indeed, as respiratory infections are time- and exposure-dependent, it is possible that differences in follow-up times and in symptom assessments (influenza-like illness, respiratory illness or COVID-19) have affected the power to detect associations. Second, while all the studies reported the numbers in the treatment and control arms, we did not have access to raw data and could not adjust the analysis by within-study variables. As discussed, we also did not have access to the raw values for secondary transmission (source control) in one of the studies [19]. As a work-around, we performed a meta-analysis including within-study adjusted odds ratios. However, this method comes with limitations of its own as because, in practice, no studies have exactly the same covariate definitions, which bias the estimates. Third, the mask types and instructions for mask use together with follow-up times varied by study, which likely increases between-study heterogeneity. We accounted for the biases through subgroup analysis by age group, setting and non-compliance in controls and meta-regression by non-compliance in controls.

### 4.4 Conclusions and future implications

This systematic review and meta-analysis using 17 randomized controlled trials across different exposure settings and age groups provides support for the public health policy of face mask use to limit the spread of infectious respiratory diseases, such as COVID-19. Our analysis suggests that face masks can decrease the probability of contracting a respiratory infection and can be particularly useful when combined with other personal protection methods.

Recommendations and clear communication about the benefits of face masks should be given by policymakers to limit the number of respiratory infections and, ultimately, deaths in respiratory disease pandemics, thus providing time for vaccine and treatment development.

## Data Availability

The authors confirm that the data supporting the findings of this study are available within the article, and the references 12-28.

## Conflict of intrest

The authors declare no conflict of interest.

## Contributorship statment

HMO and LTL conceived the study and conducted the main analysis. JK, MP, RS and AR assisted with the analyses and drafted the initial manuscript with HMO and LTL. All the authors participated in the interpretation, contributed to the drafts of the manuscript, and approved the final version. HMO and LTL are the guarantors and ensure that all the listed authors meet the authorship criteria and that no others meeting the criteria have been omitted.

## Data sharing

We provide raw numbers and adjusted odds ratios as supplementary information along with the code that was used to compute the associations.

## Appendix Study criteria

**Table A1:**
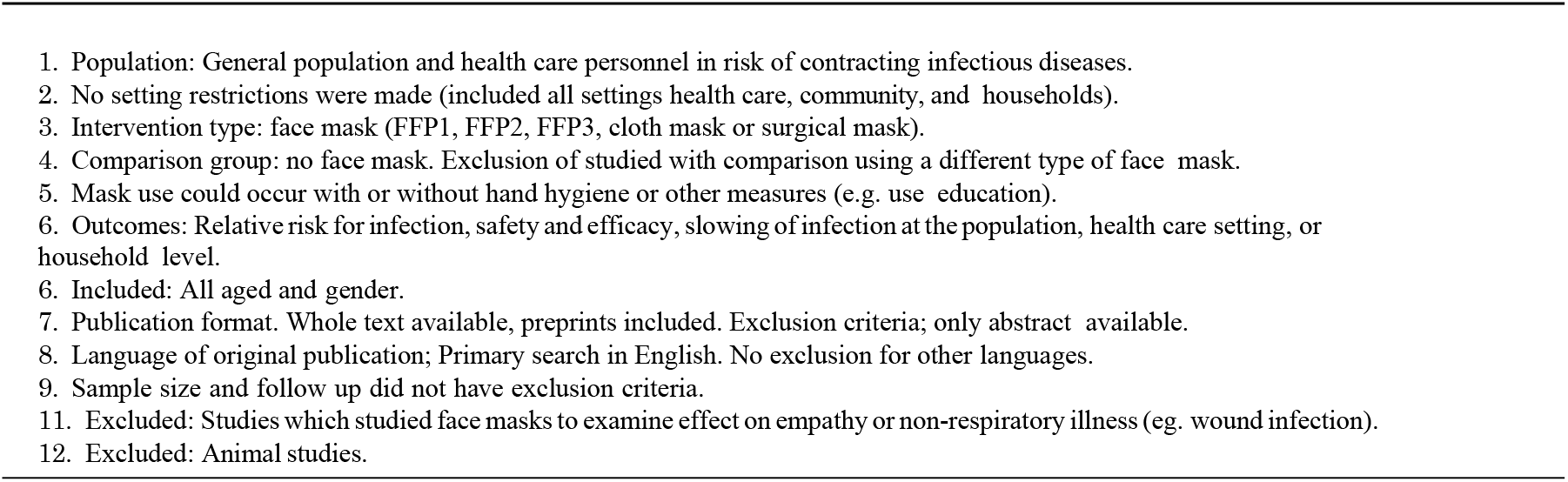
Study criteria.

### A. Search terms

Our literature search can be replicated using the following protocol.

### B. Cochrane search

**Keywords** — facemasks, infection OR “face masks”. Search with i) “facemasks, infection” resulted 47 items. Search with ii) “face masks, infection” resulted 146 items.

- Take those results found on RCT tab (add to table for flow chart).
- Compute total number from all the searches before duplicates (add to table for flow chart)
- Remove duplicates and record their number (add to table for flow chart)
- Keep only those RCT studies that measure the respiratory infection of the person wearing the mask OR those that measure protection from respiratory infections towards others.
- Do not keep articles that measure empathy or other related traits

### C. Pubmed search

**MeSH terms** — On tab *Search trial* (”Masks”[Mesh] OR “Respiratory Protective Devices”[Mesh] OR “mask” OR “facial mask”) AND (”infection”OR “Encephalitis, Viral”[Mesh] OR “Viral Load”[Mesh] OR “Central Nervous System Viral Diseases”[Mesh] OR “Influenza, Human”[Mesh] OR “Influenza A Virus, H5N1 Subtype”[Mesh] OR “Influenza A Virus, H1N1 Subtype”[Mesh] OR “Influenza A virus”[Mesh] OR “SARS Virus”[Mesh] OR “viral infection” OR “corona virus” or “swine flu” or “MERS”) OR “COVID-19” [Supplementary Concept]) AND (”Systematic review” OR “cohort study” OR “case-control” OR “Randomized Controlled Trial” [Publication Type] OR “Randomized Controlled Trials as Topic”[Mesh] OR “Controlled Clinical Trial” [Publication Type] OR “Meta-Analysis” [Publication Type] OR “Meta-Analysis as Topic”[Mesh] OR “Network Meta-Analysis”[Mesh])

### iv) Search resulted in 2,161 items (including duplicates)

- Based on abstracts of the studies from Pubmed.
- Take RCTs.
- Keep only those RCTs that measure the respiratory infection of the person wearing the mask OR those that measure protection from respiratory infections towards others.
- Do not keep articles that measure empathy or other related traits.

### D. Web of sciences

**Search terms** — Facemask AND infection AND randomized controlled trials, Face mask AND infection AND randomized controlled trials

v)Search with “Facemask AND infection AND randomized controlled trials” resulted in 63 items.

vi)Search with “Face mask AND infection AND randomized controlled trials” resulted in 13 items.

## Supplementary materials

**Figure A1:**
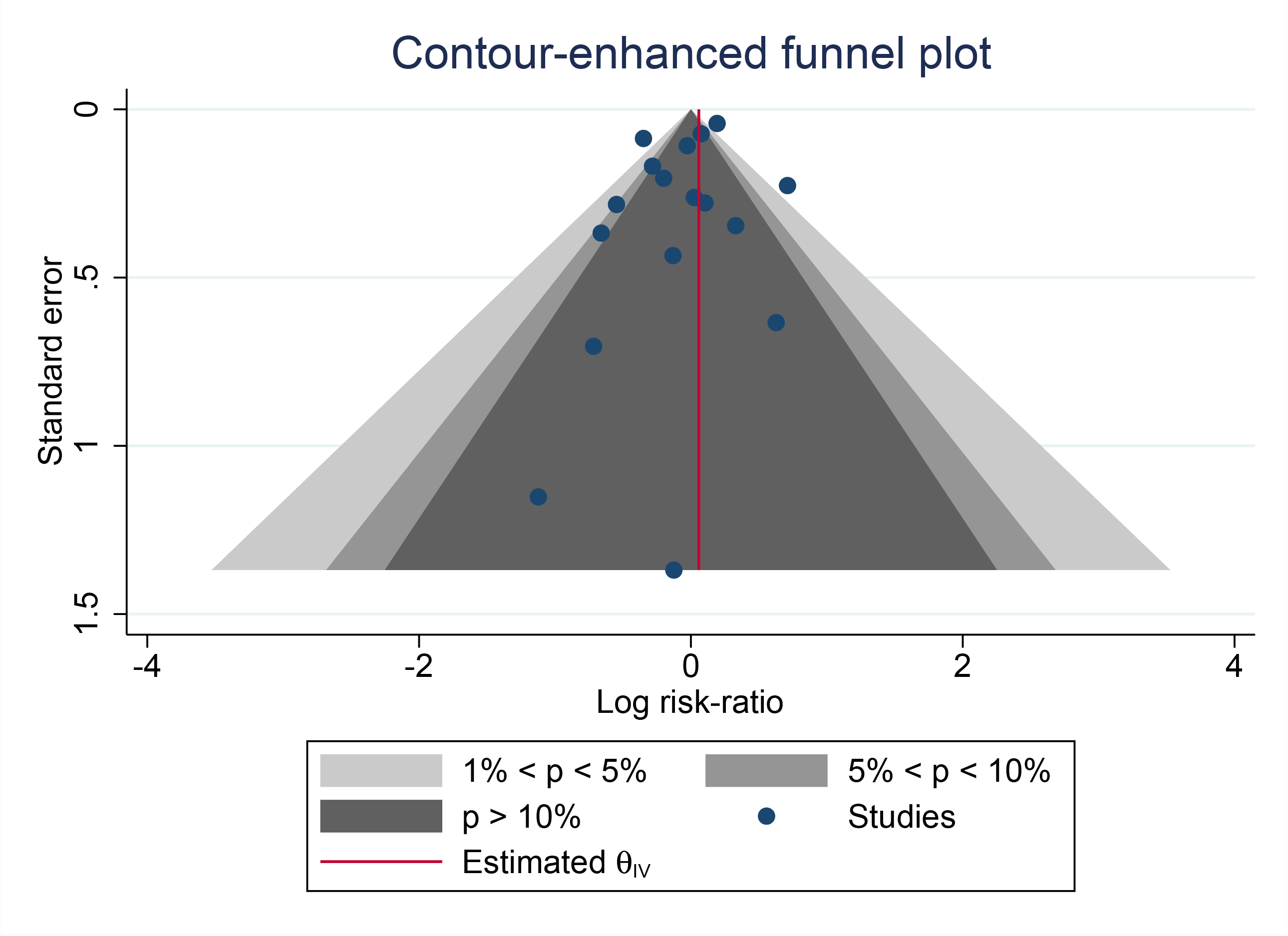
Random effects meta-analysis of the adjusted odds ratio risks of respiratory infection. The figure includes both fixed-effects and random-effect models.

**Supplementary Table.** Study characteristics R code for running analyses. Values from original articles to use alongside the R code.

PRISMA 2009 Checklist
| Section/topic | # | Checklist item | Reported on page # |
| --- | --- | --- | --- |
| **TITLE** |  |  |  |
| Title | 1 | Identify the report as a systematic review, meta-analysis, or both. | cover |
| **ABSTRACT** |  |  |  |
| Structured summary | 2 | Provide a structured summary including, as applicable: background; objectives; data sources; study eligibility criteria, participants, and interventions; study appraisal and synthesis methods; results; limitations; conclusions and implications of key findings; systematic review registration number. | abstract |
| **INTRODUCTION** |  |  |  |
| Rationale | 3 | Describe the rationale for the review in the context of what is already known. | 1 |
| Objectives | 4 | Provide an explicit statement of questions being addressed with reference to participants, interventions, comparisons, outcomes, and study design (PICOS). | 1 |
| **METHODS** |  |  |  |
| Protocol and registration | 5 | Indicate if a review protocol exists, if and where it can be accessed (e.g., Web address), and, if available, provide registration information including registration number. | 2 |
| Eligibility criteria | 6 | Specify study characteristics (e.g., PICOS, length of follow-up) and report characteristics (e.g., years considered, language, publication status) used as criteria for eligibility, giving rationale. | 2 |
| Information sources | 7 | Describe all information sources (e.g., databases with dates of coverage, contact with study authors to identify additional studies) in the search and date last searched. | 17-18 |
| Search | 8 | Present full electronic search strategy for at least one database, including any limits used, such that it could be repeated. | 17-18 |
| Study selection | 9 | State the process for selecting studies (i.e., screening, eligibility, included in systematic review, and, if applicable, included in the meta-analysis). | 2-3 |
| Data collection process | 10 | Describe method of data extraction from reports (e.g., piloted forms, independently, in duplicate) and any processes for obtaining and confirming data from investigators. | 3 |
| Data items | 11 | List and define all variables for which data were sought (e.g., PICOS, funding sources) and any assumptions and simplifications made. | 3 |
| Risk of bias in individual studies | 12 | Describe methods used for assessing risk of bias of individual studies (including specification of whether this was done at the study or outcome level), and how this information is to be used in any data synthesis. | 3 |
| Summary measures | 13 | State the principal summary measures (e.g., risk ratio, difference in means). | 2 |
| Synthesis of results | 14 | Describe the methods of handling data and combining results of studies, if done, including measures of consistency (e.g., $I^2$) for each meta-analysis. | 4 |

PRISMA 2009 Checklist
| Section/topic | # | Checklist item | Reported on page # |
| --- | --- | --- | --- |
| Risk of bias across studies | 15 | Specify any assessment of risk of bias that may affect the cumulative evidence (e.g., publication bias, selective reporting within studies). | 3-4 |
| Additional analyses | 16 | Describe methods of additional analyses (e.g., sensitivity or subgroup analyses, meta-regression), if done, indicating which were pre-specified. | 4 |
| **RESULTS** |  |  |  |
| Study selection | 17 | Give numbers of studies screened, assessed for eligibility, and included in the review, with reasons for exclusions at each stage, ideally with a flow diagram. | 5 |
| Study characteristics | 18 | For each study, present characteristics for which data were extracted (e.g., study size, PICOS, follow-up period) and provide the citations. | Table 1 |
| Risk of bias within studies | 19 | Present data on risk of bias of each study and, if available, any outcome level assessment (see item 12). | Fig 2 and 3 |
| Results of individual studies | 20 | For all outcomes considered (benefits or harms), present, for each study: (a) simple summary data for each intervention group (b) effect estimates and confidence intervals, ideally with a forest plot. | Fig 2 and 3 |
| Synthesis of results | 21 | Present results of each meta-analysis done, including confidence intervals and measures of consistency. | Fig 4 |
| Risk of bias across studies | 22 | Present results of any assessment of risk of bias across studies (see Item 15). | Fig 2 and 3 |
| Additional analysis | 23 | Give results of additional analyses, if done (e.g., sensitivity or subgroup analyses, meta-regression [see Item 16]). | 8-10 |
| **DISCUSSION** |  |  |  |
| Summary of evidence | 24 | Summarize the main findings including the strength of evidence for each main outcome; consider their relevance to key groups (e.g., healthcare providers, users, and policy makers). | 10-11 |
| Limitations | 25 | Discuss limitations at study and outcome level (e.g., risk of bias), and at review-level (e.g., incomplete retrieval of identified research, reporting bias). | 9-10 |
| Conclusions | 26 | Provide a general interpretation of the results in the context of other evidence, and implications for future research. | 11 |
| **FUNDING** |  |  |  |
| Funding | 27 | Describe sources of funding for the systematic review and other support (e.g., supply of data); role of funders for the systematic review. | cover and 11 |

From: Moher D, Liberati A, Tetzlaff J, Altman DG, The PRISMA Group (2009). Preferred Reporting Items for Systematic Reviews and Meta-Analyses: The PRISMA Statement. PLoS Med 6(7): e1000097. doi:10.1371/journal.pmed1000097
For more information, visit: www.prisma-statement.org.

